# Investigating the Causal Link Between Serum Iron Status and Pernicious Anaemia Risk: A Mendelian Randomisation Study

**DOI:** 10.1101/2024.10.28.24316258

**Authors:** Guillermo Comesaña Cimadevila, Alfie Thain, Marie-Joe Dib, Kourosh R Ahmadi

**Author notes:** Corresponding author: Kourosh R Ahmadi, School of Biosciences and Medicine, University of Surrey, Guildford, UK. Contributed equally to the manuscript (Guillermo Comesaña Cimadevila, Alfie Thain).

## Abstract

**Introduction:** Pernicious anaemia (PA) is characterised by vitamin B_12_ deficiency due to autoimmune-mediated destruction of gastric parietal cells and the consequent loss of intrinsic factor. A considerable proportion of PA patients also exhibit iron deficiency (ID), both before or at (20.7-52%) and after (46.4%) PA diagnosis. However, findings from observational studies do not clarify whether ID contributes to PA risk or is a consequence of the PA disease process. Given the high prevalence of ID at PA diagnosis, we hypothesised that reduced iron status may play a causal role in PA risk.

**Methodology:** We conducted two-sample Mendelian Randomisation (MR) analyses to evaluate the causal effect between systemic iron status and PA risk. Genetic association data for iron status were sourced from the deCODE study. Additionally, PA-relevant associative data with the chosen SNPs were obtained from genome-wide-association-study (GWAS) summary statistics, primarily from R10 *FinnGen* release and from the GWAS conducted by Laisk et al. (2021) for replication purposes. Participant data consisted of 3,694 cases of PA and 393,684 controls. Inverse-variance weighted analysis was the primary MR method, with sensitivity analyses including Egger, and weighted-median estimates, additionally to testing for horizontal pleiotropy and heterogeneity.

**Results:** Four SNPs were strongly associated with systemic iron status and were used as genetic instruments. We found that genetically predicted iron status was not significantly associated with PA risk (odds ratio per 1 standard deviation increase in serum iron: 1.13, 95% confidence interval 0.80-1.58, *P*=0.49). Sensitivity analyses had consistent results, indicating that MR assumptions were not violated and highlighting a null result subjectivity due to the presence of horizontal pleiotropy and heterogeneity.

**Conclusion:** This is the first MR study investigating the potential causal relationship between iron status and PA risk. Our results show that genetically predicted iron status is not associated with a significantly increased PA risk among individuals of European ancestry. Further research is needed to understand the manifestation of ID in PA.

## Introduction

Pernicious anaemia (PA) is a complex and life-long condition that develops during the end stages of autoimmune atrophic gastritis (AAG) (1). The disease is characterised by vitamin B_12_ (B_12_) deficiency derived from B_12_ malabsorption as a consequence of gastric atrophy leading to reduced or absent intrinsic factor production (2). The prevalence of PA is often quoted at 0.1% among the general population, rising to 1.9% among individuals aged sixty or over. However, due to frequent mis or underdiagnosis, the true prevalence is likely to be notably higher (3). The presentation of PA is most commonly with neurological and cognitive symptoms but can also present in up to 20% of patients with gastrointestinal and haematological symptoms (4). This gastrointestinal symptomology originates from an autoimmune-mediated attack on parietal cells (PCs) promoting an atrophic gastric environment (1). A hallmark of PA and AAG is the presence of parietal cell autoantibodies (PCAs) targeting the H^+^/K^+^ ATPase proton pump (5,6). These autoantibodies have been linked to gastric achlorhydria;, resulting in compromised hydrochloric acid production. Despite advances in understanding many aspects of the disease, the aetiology of PA remains largely unknown. Given its multifactorial nature, both genetic predispositions and environmental triggers have been proposed to contribute to the onset, progression, and severity of PA (7).

Iron deficiency (ID) is the most prevalent nutrient deficiency worldwide; affecting over 1 billion people, and more predominantly observed among the female population (8). Causes of ID are diverse but commonly results from inadequate dietary intake, blood loss, or impaired duodenal absorption, which if long-term and untreated, can progress to iron deficiency anaemia (IDA) (9). Among PA patients, ID is observed both before and after PA diagnosis, with previous studies finding ID in 20.7– 52% prior to or at the time of PA diagnosis, and 22.3–46.4% after diagnosis (10–12). Other studies have noted that 52% of patients with autoimmune gastritis and 17-29% of patients with PA presented with ID, though the timeline of ID development relative to PA diagnosis was not specified (13,14). Additionally, one study reported that ID developed in 41% of PA patients at some stage following the first year of vitamin B_12_ treatment (15). Among PA patients with ID, there is heterogenic presentation of anaemia, with individuals presenting with microcytic, normocytic, or macrocytic anaemia, in contrast to the common perception that PA patients only present with macrocytic megaloblastic anaemia (13,16). This heterogeneity has direct implications for the clinical management of PA, particularly in cases concurrent with ID. Currently, no routine screening or management protocols for ID in PA exist, despite the high prevalence of ID in this patient group. There is little doubt that absence of such protocols have led to sub-optimal management of PA patients and avoidable consequences in terms of symptom improvement and quality of life (17,18).

A number of observational studies have investigated the physiological link between ID and PA (see **Supplementary material, Figure 1)**. Results from these studies have been equivocal with no clear answer as to whether ID leads to an increased susceptibility to PA or if PA onset and/or commencement of B12 therapy increases the risk of ID (10–12,18,19). Given that randomised controlled trials (RCTs) to confirm whether the link between iron and PA is causal or consequential is not available and difficult to implement given the ethical and logistical challenges, the question remains unanswered. This gap in research, alongside an incomplete understanding of the pathophysiological link between ID and PA, presents a significant challenge for effective patient management.

Given that there is not sufficient data or evidence to support whether increased susceptibility to PA or if PA onset and/or commencement of B12 therapy increases the risk of ID, we purport that an alternative approach would be to use a Mendelian randomisation (MR) framework. The MR approach is now routinely used genetic epidemiological tool that leverages data from genetic variants associated with modifiable risk factors or exposures – in this current study this would be iron status – as “instruments” to test and quantify a causal link with any outcome(s), which in our case is the risk of PA (20). The approach overcomes many of the limitations of traditional epidemiological tests. Most notably, by utilising genetic variants associated with iron status as instrumental variables, rather than simply utilising iron status, MR avoides traditional confounding between the exposure of interest (iron status) and the outcome (PA).

We set out to use a MR framework as a robust and simple test of whether genetically predicted reductions in systemic iron status result in an increased risk of PA (20).

## Methods

### Study Design

A flowchart outlining the design (**Figure 1**) and the MR-specific framework (**Figure 2**) is provided for clarity. We conducted a two-sample MR analysis to test for a causal effect between iron status and risk of PA. Three key assumptions pertaining to the selection of genetic instruments were required to ensure methodological validity and to ensure the robustness of MR analyses (20). These were:

1. Independence: The genetic instruments for iron status should not be associated with risk of PA through external measured or unmeasured factors;
2. Relevance: A sufficiently robust statistical association is required between the genetic instruments used and iron status;
3. Exclusion Restriction: The variants used as instruments cannot be related to PA, through external risk factors other than the exposure of interest, namely iron status.

**Figure 1.**
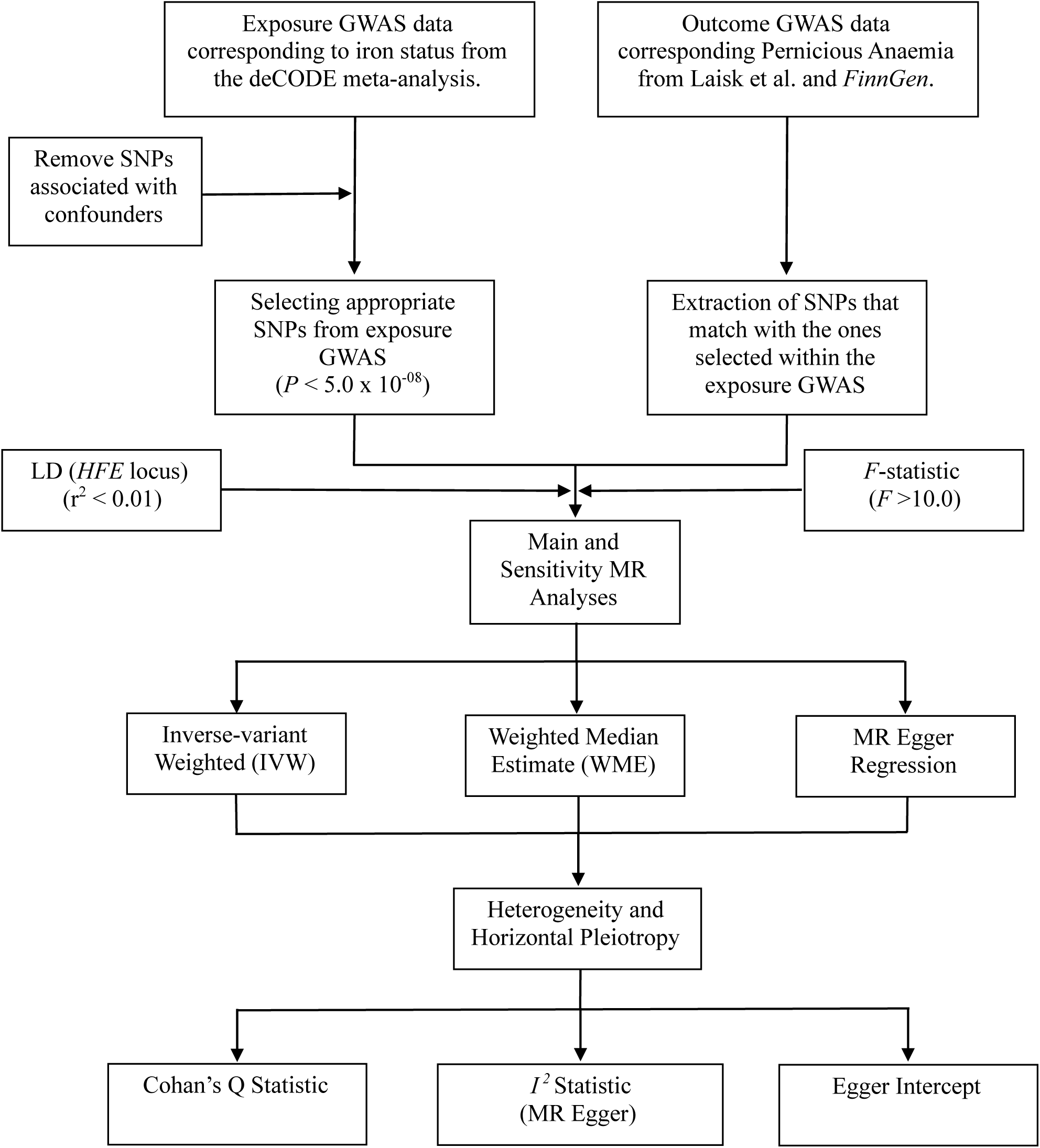
Flowchart outlining MR study protocol and study design between systemic iron status and PA. LD Coefficient = r^2^.

**Figure 2.**
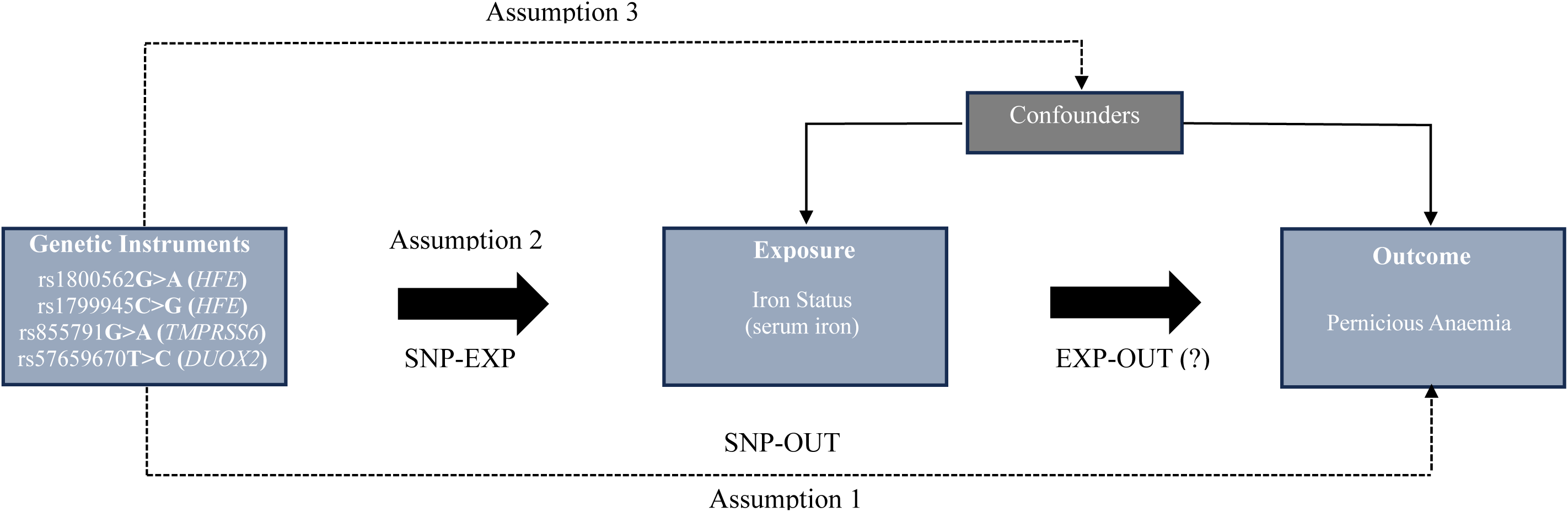
Mendelian Randomisation: Assumption 1 (Independence assumption), Assumption 2 (Relevance assumption), Assumption 3 (Exclusion restriction). SNP-Exposure association (SNP-EXP), SNP-outcome association (SNP-OUT), Exposure-Outcome association (EXP-OUT).

### Genetic Instrument Selection and Data Sources

Systemic iron status was assessed as the modifiable exposure. Various biomarkers are available for measuring intrinsic iron levels, including serum iron, transferrin saturation ratio, serum ferritin, or total iron binding capacity. To ensure robustness, the selected instrumental variants were ensured to be robustly associated with all four biomarkers, with an adjusted *P* < 5.0 x 10^-08^ threshold for statistical significance. Instrumental data was extracted from the Bell et al. (21) meta-analysis (Iceland, UK, and Denmark), composed of 3 genome-wide association studies (GWAS). Four SNPs were selected: rs1800562G>A (chr:6, exon 4, p.26092913) and rs1799945C>G (chr:6, exon 2, p.26090951) in the hereditary haemochromatosis (*HFE*) gene, rs855791G>A (chr:22, exon 17, p.37066896) in the transmembrane serine protease 6 (*TMPRSS6*) and rs57659670T>C (chr:15, exon 17, p.45106240) within the dual oxidase 2 (*DUOX2*) (21). While the SNPs located within the *TMPRSS6* and *HFE* loci have previously been found to be robustly associated with systemic iron status, the *DUOX2* locus was suggested to present a novel statistical link with iron. Overall, since multiple biomarkers would be appropriately eligible for the MR analysis, serum iron was the selected marker for iron level quantification in line with a previous publication by Hujoel et al. (22).

Given that the outcome of interest was PA, it was required to quantify its degree of association with the selected SNPs. Publicly available GWAS summary statistics from the R10 *FinnGen* release (https://r10.finngen.fi/), corresponding to data from 500,000 individuals from the Finish Biobank was the chosen primary source. The relevant data for PA were extracted by performing a local phenotypic search for “vitamin B_12_ deficiency anaemia”. The corresponding cohort comprised white individuals of Finnish descent, built by matching controls to the endpoint cases through variables including year of birth and sex. A total of 3,694 cases of B_12_ deficiency anaemia, with a mean age of 62.07 years, (50.5% being females and 49.5% males) (23). The cases were identified using the ICD-10 code for PA (D51.0), while samples with different diagnostic codes were considered controls (n=393,694).

Associations between genetic instruments and PA were also tested for replication by utilising data from British, Estonian, and Finnish descent corresponding to the GWAS conducted by Laisk et al. (24). To ensure, the selected SNPs were not subject to instrumental weakness, *F*-statistic values of instrumental strength across each individual SNP were computed. A crucial requirement of an MR study is avoiding linkage disequilibrium (LD), between the selected SNPs and other loci, as this could introduce confounding (25). As such, LD was tested across both SNPs within the *HFE* gene, by using the LDLink tool considering all populations of European descent per 1000 genomes. To ensure that MR assumption 3 (exclusion restriction) was upheld, we examined the association between the selected SNPs with well-established risk factors for PA, namely sex and age. Finally, we performed a search into each of the selected SNPs within the R10 *FinnGen* release aiming to identify possible secondary phenotypes that might influence PA, that could potentially interfere with the causal effect estimate. Therefore, further reassuring the compliance to MR assumption 1 (independence assumption).

### Mendelian Randomisation Analysis

All associations between each SNP and serum iron were combined into one arm, of which several statistical tests were conducted to examine the genetically predicted causal link between iron status and PA. The primary analytic technique was the inverse-variance weighted (IVW) meta-analysis, which combines associations between each SNP and serum iron status into a single estimate of the causal effect on PA (20). In addition, 2 sensitivity analyses were performed. First, a weighted median estimator (WME) was applied, which can provide accurate causal effect estimates even when 50% of the genetic instruments have invalid IVWs, hence minimising weak instrument bias (26). Finally, an MR-Egger regression was performed, to ensure result accuracy regardless of weak instrument bias by accounting for the presence of horizontal pleiotropy (27).

### Assessment of Heterogeneity and Horizontal Pleiotropy

It was ensured that appropriate statistical parameters were computed to account for the influence of horizontal pleiotropy and heterogeneity (28). The MR Egger intercept was included in the regression analysis as a measure of pleiotropic bias between the selected genetic instruments. Furthermore, Cochran’s Q statistic was used to assess heterogeneity between the combined effect of the selected genetic instruments on the IVW approach (29). Moreover, the *I^2^*statistic was the selected statistical test for the identification and verification of heterogeneity within the Egger regression.

### Statistical Analysis

All the analyses mentioned above were computed using RStudio (version 4.4.0). The primary package utilised consisted of the Two-Sample MR package (version 0.4.26) extracted from GitHub (https://mrcieu.github.io/TwoSampleMR/). Data outlining the association between the selected SNPs, serum iron status, and PA was displayed as betas and standard errors, with the respective effect allele frequencies independent of each SNP. The results displaying the associative link between iron status and PA were expressed as an odds ratio, with a 95% confidence interval and a significance value set at *P* < 0.05.

## Results

Four SNPs strongly associated (*P* < 5.0 x 10^08^) with systemic iron status, or serum iron were selected as instrumental variables, which include rs1799945, rs1800562 (*HFE*), rs855791 (*TMPRSS6*), and rs57659670 (*DUOX2*). Associations between the selected SNPs and all relevant iron biomarkers have been displayed in **Supplementary Tables 1 and 2**. *F* statistic values for all four instrumental variables ranged from 32.65 to 1371.74 (**Table 1**). Using the *FinnGen* data we found that the genetic instruments were not significantly associated with any known risk factors for PA (**Supplementary Table 4**). In addition, none of the selected SNPs were robustly associated with either sex or age (**Supplementary Table 5**).

**Table 1.** Characteristics of selected SNPs and associations with serum iron status and Pernicious Anaemia.

| SNP | Effect:Null | EAF | *F*-statistic | LD | Exposure (Serum Iron) |  |  | Outcome (Pernicious Anaemia) |  |  |  |
| --- | --- | --- | --- | --- | --- | --- | --- | --- | --- | --- | --- |
|  |  |  |  |  | Beta | SE | *P*-value | EAF | Beta | SE | *P*-value |
| rs1799945 | G : C | 0.137 | 853.20 | 0.006 | 0.17 | 0.0058 | 1.26 x 10<sup>-187</sup> | 0.141 | 0.032237 | 0.046 | 0.481483 |
| rs1800562 | A : G | 0.0677 | 1262.12 | 0.006 | 0.27 | 0.0076 | 1.00 x 10<sup>-200</sup> | 0.0611 | -0.01981 | 0.069 | 0.774113 |
| rs57659670 | C : T | 0.0753 | 32.65 | -- | -0.042 | 0.0074 | 1.08 x 10<sup>-08</sup> | 0.0763 | 0.01339 | 0.059 | 0.819947 |
| rs855791 | A : G | 0.431 | 1371.74 | -- | -0.17 | 0.0046 | 1.00 x 10<sup>-200</sup> | 0.418 | 0.10113 | 0.032 | 0.001450 |

**Note:** Effect Allele:Null Allele, Effect Allele Frequency (EAF), Beta = effect size, Standard Error (SE), SNP-exposure significance was set at $P < 5.0 \times 10^{-08}$ . The effect allele refers to the recessive allele within such gene, hence indicating that the wild-type allele refers to the null variant. LD Coefficient = $r^2$ , $F > 10.0$ = strong genetic instrument, $F < 10.0$ = subject to weak instrument bias. LD was calculated and extracted making use of the LDLink tool. The summary statistical data set refers to the R10 *FinnGen* release.

The IVW analysis indicated that a 1 standard deviation increase in genetically predicted serum iron levels was associated with a 13.0% higher risk of PA (OR=1.13, CI=0.80-1.58, *P*=0.49); however, the result was not statistically significant (**Table 2**). Sensitivity analyses using WME estimates supported the IVW findings, presenting a null statistical significance (OR=1.14, CI=0.88-1.48, *P*=0.32). Similarly, MR Egger estimates were also not statistically significant (OR=1.04, CI=0.30-3.59, *P*=0.95). There was no evidence of directional, horizontal pleiotropic effect given that the Egger intercept did not differ from the null (*P*=0.91). The combined heterogeneity of all the selected instruments measured with Cochran’s Q-statistic indicated no directional pleiotropy or notable heterogenic effect within the main analysis (IVW) (Q=6.54, *P*=0.051). However, a moderate-high heterogenic effect was identified to impact the Egger regression (*I^2^*=61.27%).

**Table 2.** MR estimates for the causal association between serum iron status and PA using the *FinnGen* [37] data set.

| MR Method | OR | CI | P-value | Cochran Q | Cochran Q P-value | MR Egger |  |
| --- | --- | --- | --- | --- | --- | --- | --- |
|  |  |  |  |  |  | Intercept P-value | I<sup>2</sup> (%) |
| IVW | 1.13 | 1.58 ± 0.80 | 0.492311 | 6.54 | 0.051542 |  |  |
| MR Egger | 1.04 | 3.59 ± 0.30 | 0.952501 |  |  | 0.910154 | 61.27 |
| WME | 1.14 | 1.48 ± 0.88 | 0.318990 |  |  |  |  |

**Note:** Odds Ratio (OR), 95% Confidence Interval (CI) – Upper Bound ± Lower Bound, Inverse Variance Weighted (IVW), Weighted Median Estimate (WME). Egger Intercept P-value was compared to the IVW method intercept of 0.

Replication MR using data from the GWAS data from Laisk et al. (24) revealed that per 1 standard deviation increase in genetically predicted serum iron status was associated with a non-significant lower PA risk in IVW analysis (OR=0.76, CI=0.52-1.12, *P*=0.16) (**Table 3**). WME results were similarly non-significant (OR=0.73, CI=0.52-1.05, *P*=0.087). Likewise, Egger estimates were also not statistically significant (OR=0.90, CI=0.24-3.47, *P*=0.89). No horizontal pleiotropic effect appeared to impact our findings; given that the Egger intercept did not differ from zero (*P*=0.818). Integrated heterogeneity of all the selected instruments indicated no directional pleiotropy or notable heterogenic effect within the IVW analysis (Q=6.54, *P*=0.088). Conversely, a moderate heterogenic effect was identified to impact the Egger regression (*I^2^*=54.12%).

**Table 3.** Replication of MR estimates for the causal association between serum iron status and PA, using the PA GWAS data set.

| MR Method | OR | CI | P-value | Cochran Q | Cochran Q P-value | MR Egger |  |
| --- | --- | --- | --- | --- | --- | --- | --- |
|  |  |  |  |  |  | Intercept P-value | I<sup>2</sup> (%) |
| IVW | 0.76 | 1.12 ± 0.52 | 0.163135 | 6.54 | 0.08811 |  |  |
| MR Egger | 0.90 | 3.47 ± 0.24 | 0.896621 |  |  | 0.81811 | 54.12 |
| WME | 0.73 | 1.05 ± 0.52 | 0.087432 |  |  |  |  |

**Note:** Odds Ratio (OR), 95% Confidence Interval (CI) – Upper Bound ± Lower Bound, Inverse Variance Weighted (IVW), Weighted Median Estimate (WME).

## Discussion

To our knowledge, this is the first study to directly test for a causal relationship between iron status and an increased risk of PA using an MR framework. As presented, if the MR assumptions hold, our study findings suggest no causal effect of genetically predicted systemic iron levels on PA risk. These findings align with the current consensus based on our understanding of the aetiology of AIG and PA disease process and the published literature that ID is more likely a manifestation of PA progression rather than an aetiological cause of PA (11,19).

### Does ID develop after commencement of vitamin B12 treatment among PA patients?

As previously mentioned, ID is prevalent among PA patients before, at, or after diagnosis. However, when evaluating these studies, it is important to consider the timing of ID onset, given that ID can be masked by B_12_ deficiency due to ineffective erythropoiesis and may become apparent only after the initiation of B_12_ treatment (30,31). While a causal link between ID and PA has been proposed (32), no systematic studies have thoroughly investigated the physiological basis of the association. The primary argument supporting the causal hypothesis relates to the higher prevalence of ID before PA diagnosis (10,12). Nonetheless, the early onset of ID could be subject to a different explanation due to the faster depletion of iron stores with respect to vitamin B_12_ (13,30). This will result in hepatic iron reserves being used to fulfil homeostatic demands, in light of a compromised intestinal uptake of non-haem iron due to a lack of gastric acidity as observed in AAG and PA. It is hypothesised that this manifestation would be observed before the characteristic loss of intrinsic factor and presentation of B_12_ deficiency. This has been previously presented as a potential mechanistic route that would favour the idea of a consequential relationship between iron depletion and PA development (30). It is important consider the possibility of ID being as an early-onset sign of PA for patient screening, particularly considering the delayed (>2 years on average) diagnosis of PA pertaining to the initial manifestation of the diverse symptomology associated with PA (17,33).

### Mechanistic Pathway Linking Hypochlorhydria and Iron Deficiency

The development of hypochloridria as a result of an autoimmune attack by PCAs has been proposed as a key mechanistic pathway leading to ID, followed by the development of PA (**Figure 1, supplementary material**).

In the healthy state, PCs produce both IF for B_12_ absorption and H^+^ via the H^+^/K^+^ ATPase pump to form with chloride (Cl-) to produce HCl, maintaining a gastric pH < 3.0 (34). The gastric acidic environment provided by functional parietal cells supports iron absorption by enabling the ferrireductase enzyme Duodenal Cytochrome B (DCytB) to reduce ferric iron (Fe³⁺) into its absorbable ferrous form (Fe²⁺), which is then transported into enterocytes via divalent metal transporter 1 (DMT1) (35,36). Additionally, Haem iron (from animal sources) is dissociated from dietary proteins in the acidic stomach environment and absorbed via the haem transporter (HaemT) (37). Once ferrous iron is inside the enterocyte, it is either stored as ferritin or exported into circulation as Fe²⁺ through ferroportin (FPN1), where it is oxidised back to Fe³⁺ by hephaestin (HEPH) and bound to transferrin (Tf) for systemic distribution (38,39).

In a disease state, PCAs target the H^+^/K^+^ ATPase, impairing H^+^ and IF secretion. Over time, this results in reduced HCl and IF production, leading to increased gastric pH and PA, respectively (6). Achlorhydria (pH > 5.0) compromises the acid-dependent activity of DCytB, reducing Fe³⁺ to Fe²⁺ for DMT1-mediated iron absorption (40). Furthermore, Fe³⁺ precipitation occurs in this less acidic environment, reducing the bioavailability of non-haem iron (which constitutes 80% of dietary iron). Haem iron absorption is similarly reduced due to impaired dissociation from dietary proteins, leading to decreased uptake via HaemT. As a result of these disruptions, less iron is transported out of enterocytes via FPN, and less ferric iron binds to transferrin for systemic circulation (41). This reduced iron availability impairs iron-dependent physiological processes such as erythropoiesis, macrophage iron recycling, and hepatic uptake, leading to systemic iron deficiency (42).

Advanced stages of AIG are thought to increase the likelihood of ID (43). However, Lagarde et al. found no differences in fundic atrophic damage in two groups of PA patients, either presenting low or adequate serum ferritin levels (19). A limitation of that study is that PCA measurements were not conducted. Similarly, Roguez et al. observed comparable levels of atrophic gastritis in individuals with PA and ID compared with those with PA or ID alone (11). Importantly, quantification of PCA concentration showed significantly higher concentrations of PCA among the PA+ID group compared with the PA-only group. Both studies included predominantly female patients under the age of 50 years, suggesting a certain degree of ID could have been attributed to menstrual blood loss rather than impaired duodenal absorption (44).

Further supporting the hypothesis that PA may be the preceding causal factor for ID can be extracted from studies investigating gastric juice therapy. Cook et al. demonstrated that whole gastric juice therapy, including both hydrochloric and ascorbic acid, restored a normal acidic environment (pH< 3.0), which is essential for preventing Fe^3+^ precipitation and enhancing iron absorption capacity (45). In the cohort of patients with achlorhydria, iron absorption increased more than twofold when gastric juice was administered (50.6%) compared to without it (21.8%). However, the lack of quantification to examine possible improvements in iron biomarkers presents a limitation of this study. More recently, Roguez et al. found that oral haem Fe^2+^ supplementation failed to increase serum ferritin concentration, in a small sample of PA patients with ID (n=28) (11). These results challenge the expectation and our understanding that haem Fe^2+^ form (ferrous sulfate and ferrous fumarate) is soluble in alkaline PH, does not require chelation for enterocytic uptake (46). The small sample size, with 11 out of the total 28 participants receiving oral ferrous supplementation, may explain this discrepancy, or it could relate to the uncommon presentation of a refractory response to iron supplementation (47).

Another crucial and yet overlooked micronutrient that may be playing a significant role in the iron deficiency observed among PA patients is vitamin C, an essential nutrient that must be obtained through dietary intake (48). Vitamin C is important for promoting iron absorption as it facilitates the conversion of dietary ferric iron (Fe^3+^) into the ferrous form (Fe^2+^) (49). A reduction in gastric acidity, which would be typically observed in PA, can also impair the secretion and concentration of vitamin C in the gastric juice, potentially due to a combination of reduced bioavailability, hypochloridria, and increased demand caused by its active secretion into the gastric juice.

The prevalence of vitamin C deficiency or insufficiency among PA patients is unknown although it is estimated to affect around 1.4% (2.2% of males and 0.8% of females) of the general UK population and up to 14% of the elderly (50,51). In PA patients, the loss of this micronutrient could exacerbate the already reduced iron absorption due to low gastric acid levels. Therefore, secondary vitamin C deficiency could impact iron bioavailability, and contribute to the high prevalence of iron deficiency seen at stages before, at, or after PA diagnosis.

### Can micro and macrocytosis explain an indirect relationship between PA and ID?

A common feature encountered in the literature when investigating PA and ID patients is the complexity of the type of anaemia present (11,12,14,19,30). Observations that PA patients with ID (with or without anaemia), presenting with microcytosis and/or macrocytosis have been widely reported in the published literature. Further research is needed to dissect the mechanisms linking micro and macrocytic anaemia, and PA mediated by erythropoiesis among PA patients with or without ID. It has been suggested that ID may be a result of menstrual blood loss and intestinal malabsorption playing a less important role (11,19). However, it is likely that a combination of factors, including menstrual blood loss, intestinal malabsorption, baseline iron status before AAG development and dietary iron intake, would contribute to ID development (47,52). These underlying factors could impact the rate and severity of ID development among PA patients.

Finally, the commencement or regimen of B_12_ therapy can also affect the development of iron deficiency and/or microcytic anaemia. Under this scenario ID and microcytosis manifest as a direct consequence of B12 therapy independently of a disease process. Based on this observation, we hypothesise that patients using more frequent B_12_ injections – as commonly observed – will exhibit a shift from B_12_ deficiency and/or macrocytosis to iron deficiency and/or microcytosis, due to an increased demand for iron, following increased erythropoiesis, and therefore requiring regular iron therapy (30,31). So far, researchers have directly attributed the development of ID among PA or AAG subject patients to the autoimmune-mediated attack causing the atrophic degradation of the oxyntic mucosal glands and PCs (6). Consequently, the important question of what specific PA or AAG-driven mechanism(s) cause ID remains unclear and continues to impact effective patient management (11,14,18).

### Strengths and Limitations

Strengths of our study include using a novel two-sample MR design, which, under certain assumptions, allow us to test for the first time whether there is a causal relationship between ID and PA, which is unaffected by confounding factors that would normally provide an Achilles heel in traditional observational studies. We used two independent datasets, each composed of a large sample of well-defined PA cases. However, our study has some limitations which warrant further discussion. First, our data and results are limited to individuals of European descent, restricting direct extrapolation of results to other ethnic groups, particularly those at elevated risk of ID. Secondly, the genetic instrument selection was a challenge, and in this study, instruments were chosen based on their association with all four iron status markers. While the use of four independent instruments with a clear mechanism related to iron status overcomes part of this limitation, incorporating more SNPs which capture different aetiological aspects of iron status would be a more powerful and robust approach. Despite statistical methods, including the *F*-statistic or Egger intercept suggesting a null presence of directional pleiotropy, the possibility of residual horizontal pleiotropic effects could have occurred. More specifically, the value for heterogeneity (*I^2^*) in the Egger regression analysis exceeded 50%, suggesting that results should be interpreted with moderate caution. Lastly, reliance on the ICD-10 coding for PA, which is used liberally by clinicians, is highly problematic, and could have led to misclassification of PA; i.e. individuals being wrongly labelled as having PA or not having PA. It is highly likely that true PA (D51.0) is under-diagnosed/reported whilst “anaemia” is over-diagnosed given that all 5 ICD-10 codings for B_12_ deficiency are labelled with anaemia.

### Future Work

A logical next step would be to conduct a bidirectional MR study to assess whether genetic variants associated with a higher susceptibility to PA are causally associated with lower iron status, using a novel set of PA-specific instruments. Such findings could strengthen results from observational studies linking PCA-driven reductions in gastric acidity, impairing the intestinal absorption of iron. Additionally, future research with a broader selection of ID-specific instruments should be conducted, placing the focus on ID rather than solely iron status *per se*. Further research is also needed to explore the role of B12 therapy on erythropoiesis and ID and test whether those using more frequent B12 injections exhibit higher rates of ID and/or microcytic anaemia.

Finally, future studies should also investigate the potential role of vitamin C as a mediator in this mechanism. An MR study assessing whether vitamin C deficiency is causal for PA and/or whether PA leads to vitamin C deficiency could provide important insights into this clinical picture. If vitamin C deficiency is found to be a key driver of impaired iron absorption in these patients, targeted interventions such as vitamin C supplementation could become part of clinical care. Clinical trials could also be used to assess whether vitamin C supplementation improves iron absorption in PA patients. Including vitamin C status in PA screening, particularly in patients with iron deficiency, may be beneficial. In all cases, the overarching goal would be to improve the management of PA patients through targeted treatment of ID.

## Conclusion

This study suggests that among individuals of European descent iron status is not an important driver of elevated risk of PA. However, bidirectional MR studies are important to investigate whether PA could be the underlying cause of ID. Whether ID and PA are subject to either a causal or consequential relationship should be ideally confirmed through clinical trials, with a goal of improving patient management to accomplish a universal precision medicine approach. This study lays the groundwork for further research that could improve clinical guidelines aimed at improving care for PA patients.

## Supporting information

Associations between the selected SNPs and all relevant iron biomarkers have been displayed in Supplementary Tables 1 and 2.

## Data Availability

All data produced in the present work are contained in the manuscript

