## Supplementary material for "Investigating the Causal Link Between Serum Iron Status and Pernicious Anaemia Risk: A Mendelian Randomisation Study": Associations between the selected SNPs and all relevant iron biomarkers have been displayed in Supplementary Tables 1 and 2.

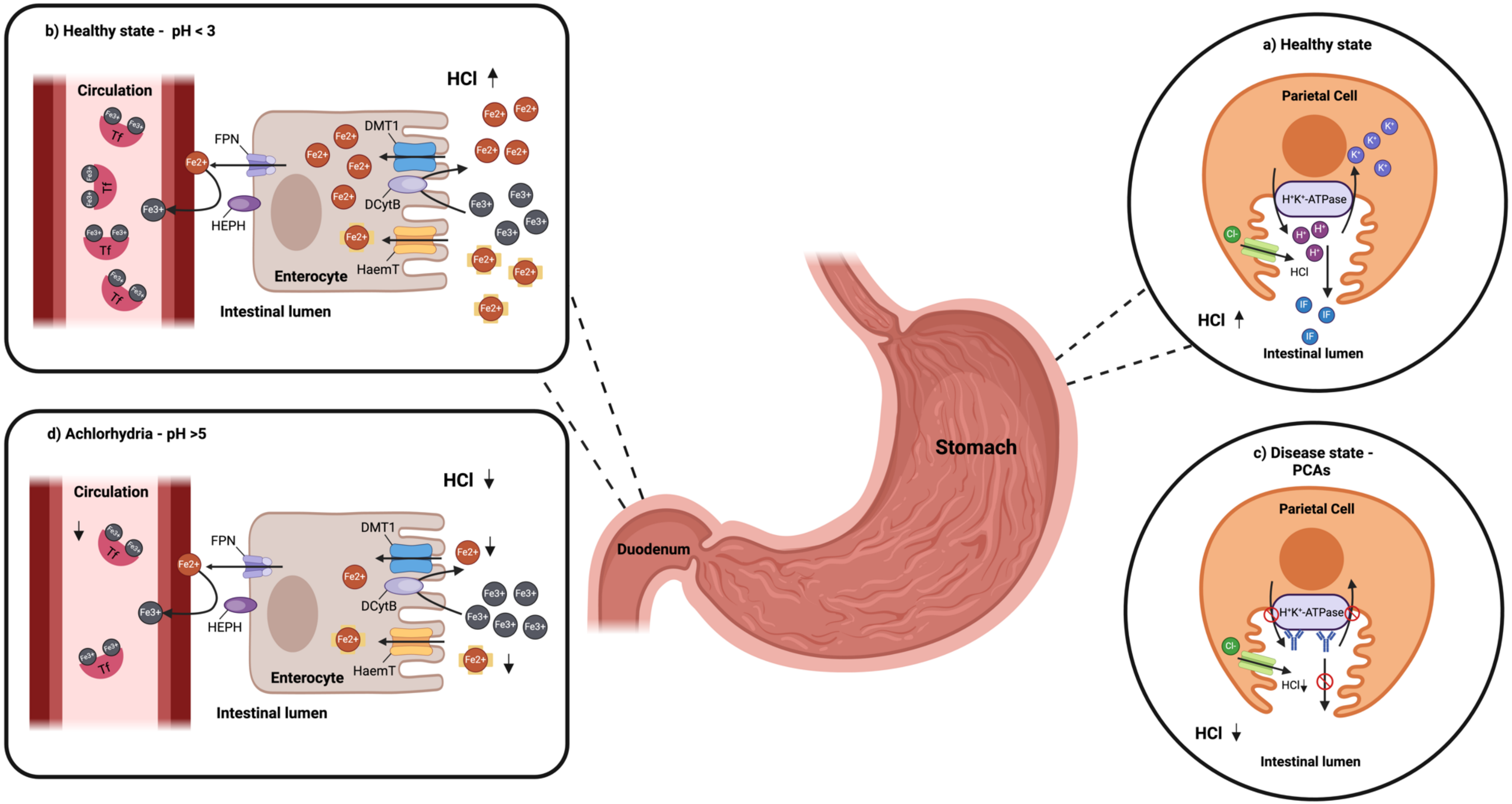


**Supplementary Figure 1.** Mechanistic pathway linking iron absorption and pernicious anaemia through parietal cell function and gastric pH changes

**Supplementary material** Methodological script written in R. The text following a "#" refers to a comment, not to any coding function or command line.

```
##Install Packages
#TwoSample MR package - documentation available at:
https://mrcieu.github.io/TwoSampleMR/

install.packages("remotes")
remotes::install_github("MRCIEU/TwoSampleMR@0.4.26")

force = TRUE

##load packages
library(TwoSampleMR)

##set working directory
setwd("/Users/guillermocomesanacimadevila/Desktop/(MSc) HUMAN NUTRITION/GENETIC
EPIDEMIOLOGY PROJECT/R DATA")

#########################################################
#set the following parameters:
#pval<- 5e-08
#r2<-0.001

#In this case, we don't have to do this because we are using a pre-selected list of
genetic instruments for iron status. If we didn't have this, we would have
#to select genetic instruments ourselves on the basis of the P-value from the GWAS
(<5E-08) and an r2 threshold of 0.001 to ensure that we are selecting independent
genetic instruments
#This is usually done through "clumping" - this will be explained in recommended
reading. Note we skip the clumping step in this code.

#Here we are pre-allocating the "results" data frame
results<-data.frame(
  exposure=character(),outcome=character(),snps=numeric(),
  ivw.OR=numeric(),ivw.l=numeric(),ivw.u=numeric(),ivw.p=numeric(),
  wm.OR=numeric(),wm.l=numeric(),wm.u=numeric(),wm.p=numeric(),
  egger.OR=numeric(),egger.l=numeric(),egger.u=numeric(),egger.p=numeric(),
  Egger.intercept.p=numeric(),Q.p=numeric(),I2=numeric(),F.statistic=numeric(),
  stringsAsFactors = F
)
```

```
# Load exposure dataset: serum iron levels - you can download the data here:
https://www.decode.com/summarydata/ #https://www.nature.com/articles/s42003-020-01575-
zhttps://www.nature.com/articles/s42003-020-01575-z
#In this case, we have already selected the genetic instruments based on published
work: https://www.ncbi.nlm.nih.gov/pmc/articles/PMC10773400/
exposure <- read.csv("/Users/guillermocomesanacimadevila/Desktop/(MSc) HUMAN
NUTRITION/GENETIC EPIDEMIOLOGY PROJECT/R DATA/iron-instruments.csv", header = TRUE)

  # We need to convert the loaded exposure dataframe into a format that the
"TwoSampleMR" package can understand and feed into the MR computation
  exp <- format_data(
    exposure,
    type = "exposure",
    snps = NULL,
    snp_col = "SNP",
    beta_col = "beta",
    se_col = "se",
    effect_allele_col = "effect_allele",
    other_allele_col = "other_allele",
    eaf_col = "eaf",
    pval_col = "p")

  # name exposure correctly
  exp$exposure <- "serum iron levels"


  # Load outcome dataset: Pernicious Anaemia - you can download the data here:
http://www.geenivaramu.ee/tools/pernicious_anemia_Laisketal2021_sumstats.gz
Paper:https://www.ncbi.nlm.nih.gov/pmc/articles/PMC8213695/
  outcome <- read.table("/Users/guillermocomesanacimadevila/Desktop/(MSc) HUMAN
NUTRITION/GENETIC EPIDEMIOLOGY PROJECT/R DATA/b12_meta_ukbb_estbb_finngen_061020.out",
header=TRUE, sep="\t")

  ##Extract Exposure SNPs from Outcome data
  outc <- outcome[outcome$rs_number %in% exp$SNP, ]

    ## Convert outcome to "TwoSample" Format
    outc <- format_data(
      dat = outc,
      type = "outcome",
      snps = outc$rs_number,
```

```
    snp_col = "rs_number",
    beta_col = "beta",
    se_col = "se",
    eaf_col = "eaf",
    effect_allele_col = "reference_allele",
    other_allele_col = "other_allele",
    pval_col = "p.value")


  ## name outcome
  outc$outcome <- "Pernicious Anaemia"

  # Harmonise data
  dat <- harmonise_data(exp, outc, action = 1)


  # Perform MR (IVW, WM, Egger)
  # F statistic using chi-squared approximation - F statistic will be explained in
recommended readings
  dat$Fstat <- dat$beta.exposure^2 / dat$se.exposure^2

  # Perform MR
  res <- mr(dat, method_list = c("mr_ivw", "mr_egger_regression",
"mr_weighted_median"))

  # MR-Egger intercept
  pleio <- mr_pleiotropy_test(dat)

  # heterogeneity test
  het <- mr_heterogeneity(dat)
  het$I2 <- ifelse((het$Q - het$Q_df ) / het$Q < 0, 0, 100*(het$Q - het$Q_df ) /
het$Q)

  # assign values in table
  results[1,1]<- "serum iron" #exposure name
  results[1,2]<-"PA Laisk et al" #outcome name
  results[1,3]<-res[1,6] #N SNPs

  results[1,4]<-exp(res[1,7]) #IVW OR
  results[1,5]<-exp(res[1,7]+qnorm(.025)*res[1,8])#IVW lower 95%
  results[1,6]<-exp(res[1,7]+qnorm(.975)*res[1,8])#IVW upper 95%
```

```
results[1,7]<-res[1,9]#IVW p-value

results[1,8]<-exp(res[3,7])#WM beta
results[1,9]<-exp(res[3,7]+qnorm(.025)*res[3,8])#WM lower 95%
results[1,10]<-exp(res[3,7]+qnorm(.975)*res[3,8])#WM upper 95%
results[1,11]<-res[3,9]#WM p-value
results[1,12]<-exp(res[2,7])#Egger beta
results[1,13]<-exp(res[2,7]+qnorm(.025)*res[2,8])#Egger lower 95%
results[1,14]<-exp(res[2,7]+qnorm(.975)*res[2,8])#Egger upper 95%
results[1,15]<-res[2,9]#Egger p-value

results[1,16]<-pleio[1,7]#Egger intercept p
results[1,17]<-het[2,8]#IVW Q statistic p
results[1,18]<-het[2,9]# I2 statistic
results[1,19]<-mean(dat$Fstat)#mean F statistic

# write results: adjust your directory accordingly
write.table(results, file = "/Users/guillermocomesanacimadevila/Desktop/(MSc) HUMAN NUTRITION/GENETIC EPIDEMIOLOGY PROJECT/R DATA/MR-results-iron-PA-Laisketal.tsv",sep = "\t", col.names = TRUE, row.names = FALSE, append = FALSE, quote = FALSE)
```

**Supplementary Table 1.** Results per SNP acquired from Bell et al.

| Rsid | Position hg38 | Min: Maj | MAF (%) | PNT | Effect in SD and (CI) | *P*-value |
|---|---|---|---|---|---|---|
| rs1799945 | chr6: 26090951 | G:C | 13.7 | Serum | 0.17 (0.16; 0.18) | 1.26e-187 |
| | | | | Ferritin | 0.059 (0.049; 0.069) | 1.51e-31 |
| | | | | TIBC | -0.12 (-0.13; -0.1) | 4.29e-66 |
| | | | | TSAT | 0.21 (0.2; 0.23) | 6.1e-229 |
| rs1800562 | chr6: 26092913 | A:G | 6.77 | Serum | 0.27 (0.26; 0.29) | 3.66e-276 |
| | | | | Ferritin | 0.13 (0.12; 0.15) | 1.85e-84 |
| | | | | TIBC | -0.45 (-0.47; -0.43) | 1e-300 |
| | | | | TSAT | 0.45 (0.42; 0.47) | 1e-300 |
| rs57659670 | chr15: 45106240 | C:T | 7.53 | Serum | -0.042 (-0.056; -0.028) | 1.08e-08 |
| | | | | Ferritin | -0.14 (-0.16; -0.13) | 1.05e-113 |
| | | | | TIBC | 0.077 (0.06; 0.094) | 3.67e-19 |
| | | | | TSAT | -0.058 (-0.074; -0.041) | 5.73e-12 |
| rs855791 | chr22: 37066896 | A:G | 43.1 | Serum | -0.17 (-0.18; -0.16) | 1e-300 |
| | | | | Ferritin | -0.044 (-0.051; -0.038) | 6.14e-37 |
| | | | | TIBC | 0.026 (0.017; 0.035) | 2.88e-08 |
| | | | | TSAT | -0.17 (-0.18; -0.16) | 1e-300 |

**Note:** Minor:Major Allele (Min:Maj), Serum Iron (Serum), Serum Ferritin (Ferritin), Total Iron Binding Capacity (TIBC), Transferrin Saturation (TSAT), 95% Confidence Interval (CI), Standard Deviation (SD), Phenotype (PNT).

**Supplementary Table 2.** Genetic instrument strength.

| | Effect on Exposure (units of SD) | | | | |
|---|---|---|---|---|---|
| | β Estimate | SE | *P*-value | $R^2$ | *F*-statistic |
| Serum | | | | | |
| rs1799945 | 0.17 | 0.0058 | 1.26e-187 | 0.68 | 1125.08 |
| rs1800562 | 0.27 | 0.0076 | 3.66e-276 | 0.92 | 1518.66 |
| rs57659670 | -0.042 | 0.0073 | 1.08e-08 | 0.02 | 40.18 |
| rs855791 | -0.17 | 0.0046 | 1e-300 | 1.42 | 2351.05 |
| Ferritin | | | | | |
| rs1799945 | 0.059 | 0.005 | 1.51e-31 | 0.08 | 202.77 |
| rs1800562 | 0.13 | 0.0067 | 1.85e-84 | 0.21 | 526.22 |
| rs57659670 | -0.14 | 0.0062 | 1.05e-113 | 0.27 | 673.67 |
| rs855791 | -0.044 | 0.0035 | 6.14e-37 | 0.09 | 233.95 |
| TIBC | | | | | |
| rs1799945 | -0.12 | 0.007 | 4.29e-66 | 0.34 | 462.72 |
| rs1800562 | -0.45 | 0.012 | 1e-300 | 2.56 | 3552.69 |
| rs57659670 | 0.077 | 0.0086 | 3.67e-19 | 0.08 | 111.91 |
| rs855791 | 0.026 | 0.0047 | 2.88e-08 | 0.03 | 44.92 |
| TSAT | | | | | |
| rs1799945 | 0.21 | 0.0065 | 6.1e-229 | 1.04 | 1385.41 |
| rs1800562 | 0.45 | 0.012 | 1e-300 | 2.56 | 3448.83 |
| rs57659670 | -0.058 | 0.0084 | 5.73e-12 | 0.05 | 61.62 |
| rs855791 | -0.17 | 0.0046 | 1e-300 | 1.42 | 1890.36 |

**Note:** For all 4 iron biomarkers, the amount of variance explained by each instrument as well as the *F*-statistic is computed. As effect sizes are in units of standard deviation, the percentage of variation in iron biomarker explained by the SNP ($R^2$); computed based on the formula; $R^2 = 2 \times \beta^2 \times AF \times (1 - AF)$, where AF = Allele Frequency.

**Supplementary Table 3.** Reported exposure and outcome associations.

| Exposure (serum iron status) | | | | | | |
|---|---|---|---|---|---|---|
| SNP | Effect Allele | Other Allele | EAF | Beta | SE | *P*-value |
| rs1799945 | **G** | C | 0.137 | 0.17 | 0.0058 | 1.26e-187 |
| rs1800562 | **A** | G | 0.0677 | 0.27 | 0.0076 | 1.0e-200 |
| rs57659670 | **C** | T | 0.0753 | -0.042 | 0.0074 | 1.08e-08 |
| rs855791 | **A** | G | 0.431 | -0.17 | 0.0046 | 1.0e-200 |
| Outcome (Pernicious Anaemia) | | | | | | |
| rs1799945 | **G** | C | 0.141 | 0.032237 | 0.046 | 0.481483 |
| rs1800562 | **A** | G | 0.0611 | -0.01981 | 0.069 | 0.774113 |
| rs57659670 | **C** | T | 0.0763 | 0.01339 | 0.059 | 0.819947 |
| rs855791 | **A** | G | 0.418 | 0.10113 | 0.032 | 0.001450 |

**Note**: Odds Ratio (OR), Beta = Effect Size (units of standard deviation for the exposure; $\log_{10}$ OR for the outcome value, Standard Error (SE). Effect Allele Frequency (EAF).

**Supplementary Table 4.** Phenotypes associated with SNPs determined through the R10 *FinnGen* release.

| SNP | Associated Phenotypes |
|---|---|
| rs1799945 | Endocrine, nutritional and metabolic diseases; disorders of iron metabolism, diseases of the circulatory system; hypertension, quantitative endpoints; height, inverse-rank normalized, drug purchase endpoints; antihypertensive medication, cardiometabolic endpoints; cardiovascular diseases (excluding rheumatic etc), diseases of the circulatory system; varicose veins, diseases of the circulatory system; atrial fibrillation and flutter with reimbursement, diseases of the circulatory system; atrial fibrillation and flutter, rheuma endpoints; gout, strict definition, cardiometabolic endpoints; other heart diseases, cardiometabolic endpoints; heart failure and antihypertensive medication, diseases of the genitourinary system; other diseases of urinary system, diseases of the circulatory system; other disorders of veins, quantitative endpoints; weight, inverse-rank normalized. |
| rs1800562 | Endocrine, nutritional and metabolic diseases; disorders of iron metabolism, diseases of the circulatory system ; hypertension, quantitative endpoints; height, inverse-rank normalized, drug purchase endpoints; antihypertensive medication , cardiometabolic endpoints; cardiovascular diseases (excluding rheumatic etc), diseases of the circulatory system; varicose veins, diseases of the circulatory system; atrial fibrillation and flutter with reimbursement, diseases of the circulatory system; atrial fibrillation and flutter, rheuma endpoints; gout, strict definition, cardiometabolic endpoints; other heart diseases, cardiometabolic endpoints; heart failure and antihypertensive medication, diseases of the genitourinary system; other diseases of urinary system, diseases of the circulatory system; other disorders of veins, quantitative endpoints; weight, inverse-rank normalized. |
| rs57659670 | Diseases of the blood and blood-forming organs and certain disorders involving the immune mechanism; iron deficiency anaemia, diseases of the blood and blood-forming organs and certain disorders involving the immune mechanism; other and unspecified iron deficiency, interstitial lung disease endpoints; diseases of the respiratory system, acute upper respiratory infection; asthma and related endpoints, bronchitis; diseases of |

| | |
|---|---|
| | the respiratory system, acute upper respiratory infections of multiple and unspecified sites, diseases of the blood and blood-forming organs and certain disorders involving the immune mechanism, diseases of the circulatory system; sequelae of cerebrovascular disease, diseases of the respiratory system; influenza and pneumonia, pregnancy, childbirth and the puerperium; other maternal diseases classifiable elsewhere but complicating pregnancy, childbirth and the puerperium, certain infectious and parasitic diseases; viral agents as the cause of diseases classified to other chapters, neoplasms, from cancer register, malignant neoplasm of colon (controls excluding all cancers), neoplasms, from cancer register (ICD-O-3), colon adenocarcinoma (controls excluding all cancers), neoplasms, from cancer register, colorectal cancer (controls excluding all cancers), neoplasms, from cancer register, colorectal adenocarcinoma (controls excluding all cancers), comorbidities of neurological endpoints, depression medications, diseases of the respiratory system; all pneumoniae, mental and behavioural disorders; (other neurotic disorders, diseases of the respiratory system; all influenza, rheuma endpoints, postmenopausal osteoporosis with pathological fracture, endocrine, nutritional and metabolic diseases, disorders of puberty, psychiatric endpoints from Katri Räikkönen ; any mental disorder, diseases of the respiratory system; acterial pneumoniae, diseases of the genitourinary system; calculus of lower urinary tract, diseases of the skin and subcutaneous tissue; erythema nodosum, endocrine, nutritional and metabolic diseases; disorders of puberty, other/unspecified, diseases of the musculoskeletal system and connective tissue; other intervertebral disc disorders, diseases of the ear and mastoid process; sudden idiopathic hearing loss, neoplasms from hospital discharges; benign lipomatous neoplasm of skin and subcutaneous tissue of head, face and neck, diseases of the genitourinary system; neuromuscular dysfunction of bladder, endocrine, nutritional and metabolic diseases; disorders of sphingolipid metabolism and other lipid storage disorders, neoplasms from hospital discharges; carcinoma in situ of colon (controls excluding all cancers), codes for special purposes Covid-19 confirmed case, diseases of the blood and blood-forming organs and certain disorders involving the immune mechanism , selective deficiency of immunoglobulin A [IgA]. |

| | |
|---|---|
| rs855791 | Diseases of the blood and blood-forming organs and certain disorders involving the immune mechanism; other and unspecified anaemias, diseases of the eye and adnexa; paralytic strabismus. |

**Supplementary Table 5.** Association of genetic instruments with sex and age.

| | Exposure | | Outcome | | | |
|---|---|---|---|---|---|---|
| SNP | Effect:Other | AF | Variant | AF | Sex *P*-value | Age *P*-value |
| rs1799945 | G : C | 0.137 | 6:26091179:C>G | 0.141 | 0.695 | 0.732 |
| rs1800562 | A : G | 0.0677 | 6:26093141:G>A | 0.0611 | 0.852 | 0.248 |
| rs57659670 | C : T | 0.0753 | 15:45398438:T>C | 0.0763 | 0.556 | 0.570 |
| rs855791 | A : G | 0.431 | 22:37462936:A>G | 0.418 | 0.107 | 0.727 |

**Note:** Variant identifier has been set in the form "chr:pos:dominant:recessive", where the effect allele is the recessive variant. Allele Frequency (AF).
